# Somatosensory differences between symptomatic and non-symptomatic retired contact sport athletes

**DOI:** 10.1101/2024.09.01.24312914

**Authors:** Brigitte Beck, Dawson J. Kidgell, Doug A. King, Ashlyn K. Frazer, Mark Tommerdahl, Alan J. Pearce

## Abstract

**Aim of the study:** Interest continues investigating pathophysiology of athlete cohorts with a history involving cumulative career exposure of repeated concussion and non-concussion impacts. One area yet to be explored involves the somatosensory system. Using a novel sensorimotor technique, this study measured the somatosensory system in retired contact sport athletes exposed to repetitive neurotrauma.

**Materials and Methods:** Retired athletes (n=85, mean age 48.6 ± 10.6 years, all male) completed a self-report survey on their playing career, number of concussions, and continuing symptoms. Participants completed somatosensory vibrotactile tasks assessing reaction time, amplitude discrimination (sequential, simultaneous), and temporal discrimination (temporal order judgment, duration discrimination). Participants were divided between those reporting persistent symptoms (“symptomatic”, n=63) and those without concerns (“non-symptomatic”, n=22).

**Results:** “Symptomatic” participants scored higher symptom scores compared to the “non-symptomatic” group (P<0.001). No differences were found between groups for age (P=0.152), number of concussions (P=0.193), total years played (P=0.385), or professional career length (P=0.711). “Symptomatic” group reaction times were slower to the “non-symptomatic” group (P<0.001). Reaction time variability were greater in the “symptomatic” group (P=0.002). Differences between groups were found for amplitude discrimination (sequential: P=0.031; simultaneous: P=0.036) and temporal order judgment (P=0.032). Significant correlations were found between total symptom scores and all somatosensory tasks. Correlations showed associations between total exposure years with temporal order judgement and reaction time.

**Conclusions:** This novel study showed altered sensorimotor perception in retired athletes with persistent symptoms. Our data adds to the growing pathophysiological evidence in those who experience repetitive neurotrauma during their playing careers.

## Introduction

Well described in sports such as boxing since the early 20^th^ century (Martland 1928), it has only been in the last two decades that attention has increased on the cumulative exposure effects of mild brain injuries (concussive and non-concussive). Recent reports of cognitive impairments, mental health, and neurodegenerative disease (Buckland et al. 2022; Lennon et al. 2023) repetitive physical neurotrauma to the brain is now recognised as a risk factor for chronic pathophysiology and neuropathology (Nowinski et al. 2022; Pearce et al. 2023).

Over recent years, there has been increasing attention on understanding the underlying pathophysiology of cohorts engaged in activities that involve repetitive neurotrauma to the brain. Studies using advanced neuroimaging, for instance diffusion tensor imaging (DTI) have shown white matter abnormalities (Hellewell et al. 2020), while neurophysiology techniques such as transcranial magnetic stimulation (TMS), have shown pathophysiology in the brains of contact sport athletes when compared to age-matched controls (Pearce et al. 2014; Pearce et al. 2018; Pearce et al. 2023).

Conversely somatosensory testing has a long history in understanding cortical function, providing potential for its value to contribute to this area of research. While somatosensory testing has been utilised in acute concussion settings (i.e. recovery from baseline (Francisco et al. 2015; Tommerdahl et al. 2016)), somatosensory testing has not yet been fully explored in those with chronic concerns with repetitive brain injuries many years after retiring from their sport.

Somatosensory assessments involve different vibrotactile patterns targeting unique cortical mechanisms, with studies reporting neural correlates of somatosensory cortical function (Tannan et al. 2007; Kerchner et al. 2012; Puts et al. 2013). For example, tactile reaction time (RT), measuring cognitive attention and sensorimotor pathways, has been closely linked to white matter structure and γ-aminobutyric acid (GABA) concentrations (Kerchner et al. 2012; Puts et al. 2013). Similarly, discrimination of vibrational amplitude (called amplitude discrimination, AD) between two stimuli applied to adjacent digits engages lateral inhibition to separate the response functions within the corresponding cortical regions (Puts et al. 2013). In healthy populations when the stimulus was applied simultaneously, AD responses were improved demonstrating an adaptation effect (Tannan et al. 2007). Temporal Order Judgment (TOJ) refers to the ability to detect the sequential timing order of two presented stimuli. In humans, the primary somatosensory cortex provides information about the locations of the two tactile stimuli. However, it remains unclear which other cortical areas are involved in processing TOJ. These areas are suspected to involve fronto-striatal pathways (Zhang et al. 2011; Favorov et al. 2019). Further, contributions from different cortical or subcortical areas may suggest the complexity of tactile TOJ, along with GABA mediated inhibition (Lee et al. 2013). Duration discrimination (DD) is based upon the smallest time duration difference perceived between two stimuli. DD has been reported to be impaired following mild and more severe traumatic brain injuries, reflecting impaired neuron-glial interaction (Francisco et al. 2015) as well as cerebellar-cortical interactions (Potts et al. 2009).

Therefore, based on the ability of somatosensory system to detect neurological changes, it is plausible that somatosensory testing is sensitive to those who are neurologically compromised (Francisco et al. 2015). Our hypothesis posits that within a group of retired athletes from contact sports, who have a history of neurotrauma exposure, somatosensory testing could differentiate between individuals who report persistent concerns and those who do not report any concerns. Secondary hypotheses included correlations between somatosensory performance with the number of concussions, and also the length of exposure years playing contact sports.

## Materials and Methods

### Participants

Participants (n = 85, male; see **Table 1** for descriptive data) were recruited and pre-screened for suitability. Pre-screening criteria included being between the age of 30-70 years, played contact sport, including at the professional levels, and sustained at least one concussion during their playing career, have a minimum of five years retirement from contact sports, have no serious neurological diseases or psychiatric disorders, and be free of upper limb musculoskeletal injury. All procedures were approved by the La Trobe University Human Research Ethics Committee (HEC18005), and participants provided written informed consent prior to data collection.

**Table 1.** Participants age, number of concussions, time since last concussion, career playing Australian football and participation in the Australian Football League, and time retired playing football. Data displayed as mean (SD).

|  | Age<br>(years) | Reported<br>concussions<br>(number) | Last<br>concussion<br>(years) | Age started<br>(years) | Age finished<br>(years) | Total Career<br>(years) | Professional Career<br>(years) | Time retired<br>(years) |
| --- | --- | --- | --- | --- | --- | --- | --- | --- |
| All<br>(n=85) | 48.6<br>(10.6) | 7.7<br>(4.4) | 19.0<br>(10.4) | 9.9<br>(2.5) | 31.1<br>(4.9) | 21.1<br>(5.9) | 9.1<br>(4.5) | 17.6<br>(9.8) |
| Symptomatic<br>(n=63) | 47.8<br>(10.4) | 8.0<br>(4.6) | 18.1<br>(10.1) | 10.0<br>(2.4) | 31.3<br>(5.2) | 21.3<br>(6.2) | 9.2<br>(4.4) | 16.7<br>(9.3) |
| Non-symptomatic<br>(n=22) | 51.0<br>(10.9) | 6.5<br>(3.8) | 21.6<br>(11.2) | 9.7<br>(2.7) | 30.2<br>(30.0) | 20.8<br>(5.5) | 8.8<br>(4.6) | 20.8<br>(10.9) |

### Symptom Self-Report

Participants reported on their playing history including age started, age they retired from their sport (which may include playing club level after retiring from their professional career), and the number of years they played professionally in their respective leagues. They also reported the number of diagnosed concussions from their club doctor, and the last diagnosed concussion (**Table 1**). Participants completed the Fatigue and Related Symptom Scale (Johansson et al. 2009; Johansson and Rönnbäck 2014) quantifying mental fatigue and other related symptoms affecting daily activities. Responses to 15 questions were answered using Likert rating scales in 0.5 steps, ranging from 0 to 3 indicated the severity (higher the number the greater the severity) experienced by the participant over the previous four weeks.

Johansson and Rönnbäck (2014) have previously reported a total score of 10.5 in the clinical TBI group was statistically significantly deviating from the control sample and was also above the 99^th^ percentile for the control group. Consequently, the authors recommended a cut-off score at 10.5. However they have also provided a caveat suggesting that participants above 10.5 may not always illustrate serious concerns with activities of daily living (Johansson and Rönnbäck 2014). Therefore, to increase the sample size of those with a history of neurotrauma but who themselves described that they were not seriously affected, we chose an arbitrary cut-off score of 14, reflecting previous research using TMS (Pearce et al. 2021).

### Somatosensory assessment

Somatosensory assessment was conducted using a two-digit vibrotactile stimulator (Cortical Metrics, North Carolina, USA). The portable device consists of two probes, approximately 5mm in diameter, delivering light vibrations at frequencies of 25-50 Hz to the participant’s index (D2) and middle (D3) digits of their non-dominant hand, while providing responses using a computer mouse with their dominant hand (Tommerdahl et al. 2016).

Testing consisted of a battery of four components assessing the ability of the participant to distinguish sensory stimuli based upon timing, frequency, amplitude and duration. Individual test batteries included simple RT, sequential AD (seqAD) and simultaneous AD (simAD), TOJ and DD. Participants were fully familiarised before data collection and were verbally instructed to respond as quickly as possible. No visual stimulus was provided during any of the testing. Apart from the familiarisation stage, no feedback of performance was provided during testing. Total time of testing was approximately 20 minutes.

Simple RT involved the participant responding as quickly as possible by clicking the computer mouse following a single pulse stimulus through the somatosensory device (Pearce et al. 2020). Reaction time involved taking both the response time and measuring the variability of those responses, and was completed at the start and completion of the battery to detect fatigue effects (slowing in RT and greater RT variability; (Tommerdahl et al. 2019). Amplitude discrimination required the participant to detect which vibration stimulus was ‘stronger’ in perception. In seqAD, the control and test stimuli were delivered one after the other; while simAD, the control and test stimuli were delivered at the same time. For both variations of the test, participants responded by choosing D2 or D3 for the ‘stronger stimulus’. TOJ involved the participant to determine which finger (D2 or D3) sensed the first of two brief pulses. DD required the participant to identify which digit perceived the stimulus delivery time was longer in duration. For all testing 20 trials were provided, except reaction time which was 10 trials at the beginning and 10 trials at the end of the battery.

### Data and statistical analyses

Scores from the Fatigue and Related Symptom Scale were summed to give a total out of a maximum score of 44. Based on the symptom self-report, comparison of participants was divided into Symptomatic (scores >14; N=63) or Non-symptomatic (scores ≤ 14; N=22) groups.

Reaction time (ms) was determined by removing the best and worst time and averaging the remaining response times. Variability in reaction times (RTvar) was the standard deviation of the mean reaction time. Amplitude discrimination was the minimum threshold (μm) that could be detected between the test and control stimuli. Temporal order judgement was calculated as the shortest interstimulus duration (ms) that could be detected by the participant. Duration discrimination was calculated as the shortest time (ms) that could be identified between the test and control stimuli. For more detailed analytics of the test batteries, see Favorov et al. (2019) and Zhang et al. (2011).

Statistical analyses were conducted using Jamovi (Jamovi Project V2.3, Sydney, Australia). Shapiro-Wilk tests were performed on the data and showed non-normal distributions (W= 0.732 – 0.957; P<0.001). Logarithmic transformations were performed, which also showed non-normal distributions (W= 0.685 – 0.861; P<0.01) and, therefore, non-parametric statistical measures were used. Mann-Whitney U tests were performed to measure the median differences between the Symptomatic and Non-symptomatic groups for self-reported fatigue and related symptom scores, the number of concussions and years exposed (total and professional), and sensorimotor response task results. For repeated measures of reaction time and reaction time variability within-between groups, a Scheirer-Ray-Hare test was completed. Cohen’s d effect sizes (<0.2=small; 0.2-0.8=moderate; >0.8=large) were utilised to compare the magnitude of differences between groups (Cohen 1988).

Spearman correlations investigated the relationships between symptom scores, the number of concussions and years exposed (total and professional) to reaction time, reaction time variability, amplitude discrimination, temporal order judgement, and duration discrimination. Interpretations of correlations utilised the following descriptive criteria: <0.2=weak, 0.2-<0.4=moderate, 0.4-<0.6=strong, and 0.8-1.0=very strong (Evans 1996). Correlations for comparisons between groups were completed by transforming the correlation coefficient into z scores using the methods described by Cohen et al. (2003). Alpha was set to P<0.05 for all tests.

## Results

All participants completed testing with no adverse effects. Between group testing showed no differences in age (U=537, P=0.152), number of self-reported concussions (U=552, P=0.193), the time since last concussion (U=543, P=0.164), total years played (U=596, P=0.385), professional career length (U=645, P=0.711), or time since retirement (U=514, P=0.092; **Table 1**).

### Comparison between groups

Between groups analysis in **Table 2** showed that the symptomatic group recorded significantly higher self-reported scores compared to the non-symptomatic group (P<0.001). Reaction time at the start of the testing battery (RT1) and at the completion of the battery (RT2) were significantly different between groups. Consequently, mean reaction times were significantly different (mean difference 110.6 ± 108.2 ms, P<0.001, d=1.0). However, no interaction for RT was seen between time and groups (H=0.52, P=0.47). Variability in reaction times at the start (RTvar 1) and completion (RTvar 2) were significantly different (**Table 2**), and mean RTvar differences observed between groups (mean difference 16.5 ± 19.2 ms, P=0.002, d=0.8). Similarly, no interaction was found between time and groups for RTvar (H=0.27, P=0.87).

**Table 2.** Self-reported fatigue and related symptom scale and somatosensory mean (SD) and median data, Mann-Whitney statistic and significance.

|  | Symptomatic (n=63) |  | Non-symptomatic (n=22) |  | Cohen's d | Mann-Whitney U | Alpha |
| --- | --- | --- | --- | --- | --- | --- | --- |
|  | Mean (SD) | Median | Mean (SD) | Median |  |  |  |
| **Total SS (max 44)** | 21.9 (5.1) | 22.0 | 8.3 (7.5) | 7.5 | 2.8 | 0 | <0.001 |
| **RT1 (ms)** | 356.3 (154.5) | 344.5 | 254.7 (42.0) | 243.8 | 1.1 | 216 | <0.001 |
| **RT2 (ms)** | 358.2 (105.2) | 317 | 238.6 (39.5) | 241.4 | 0.9 | 281 | <0.001 |
| **Mean RT (ms)** | 357.3 (122.5) | 332.1 | 246.6 (33.0) | 226.1 | 1.0 | 299 | <0.001 |
| **RTvar1 (ms)** | 38.6 (19.8) | 36.6 | 24.6 (15.2) | 21.2 | 0.7 | 354 | 0.001 |
| **RTvar2 (ms)** | 43.2 (31.0) | 33.8 | 24.9 (24.1) | 19.3 | 0.6 | 296 | <0.001 |
| **Mean RTvar (ms)** | 40.9 (21.9) | 39.0 | 24.7 (15.5) | 18.8 | 0.8 | 299 | <0.001 |
| **SeqAD (µm)** | 57.8 (28.8) | 51.5 | 41.2 (24.2) | 40.0 | 0.6 | 459 | 0.030 |
| **SimAD (µm)** | 103.5 (75.1) | 92.5 | 65.5 (47.6) | 52.0 | 0.5 | 466 | 0.036 |
| **Mean AD (µm)** | 80.7 (41.5) | 73.0 | 53.4 (29.1) | 44.5 | 0.7 | 399 | 0.005 |
| **TOJ (ms)** | 51.1 (29.4) | 41.5 | 37.3 (20.7) | 30.7 | 0.5 | 461 | 0.032 |
| **DD (ms)** | 92.6 (45.9) | 82.5 | 79.0 (30.1) | 85.0 | 0.3 | 594 | 0.429 |

SD = Standard Deviation; SS = Symptom Score; RT = Reaction Time; RTVar = Reaction Time Variability; SeqAD = sequential Amplitude Discrimination; SimAD = simultaneous Amplitude Discrimination; AD = Amplitude Discrimination; TOJ = Temporal order Discrimination; DD = Duration Discrimination; ms = milliseconds; µm = micron.

Significant differences were found between seqAD (P=0.031) and simAD (P=0.036). Similarly, TOJ showed significant differences between groups (P=0.032). No differences between groups were observed for DD (P=0.429).

### Correlations: all participants

Correlation analyses across all participants (**figure 1a – e**) showed significant associations between total fatigue and related symptoms scores and mean RT (rho=0.563; P<0.001, **figure 1a**), mean RTvar (rho=0.477; P<0.001, **figure 1b**), mean AD (rho=0.395; P<0.001, **figure 1c**), TOJ (rho=0.306; P=0.004, **figure 1d**) and DD (rho=0.231; P=0.033, **figure 1e**).

**Figure 1.**
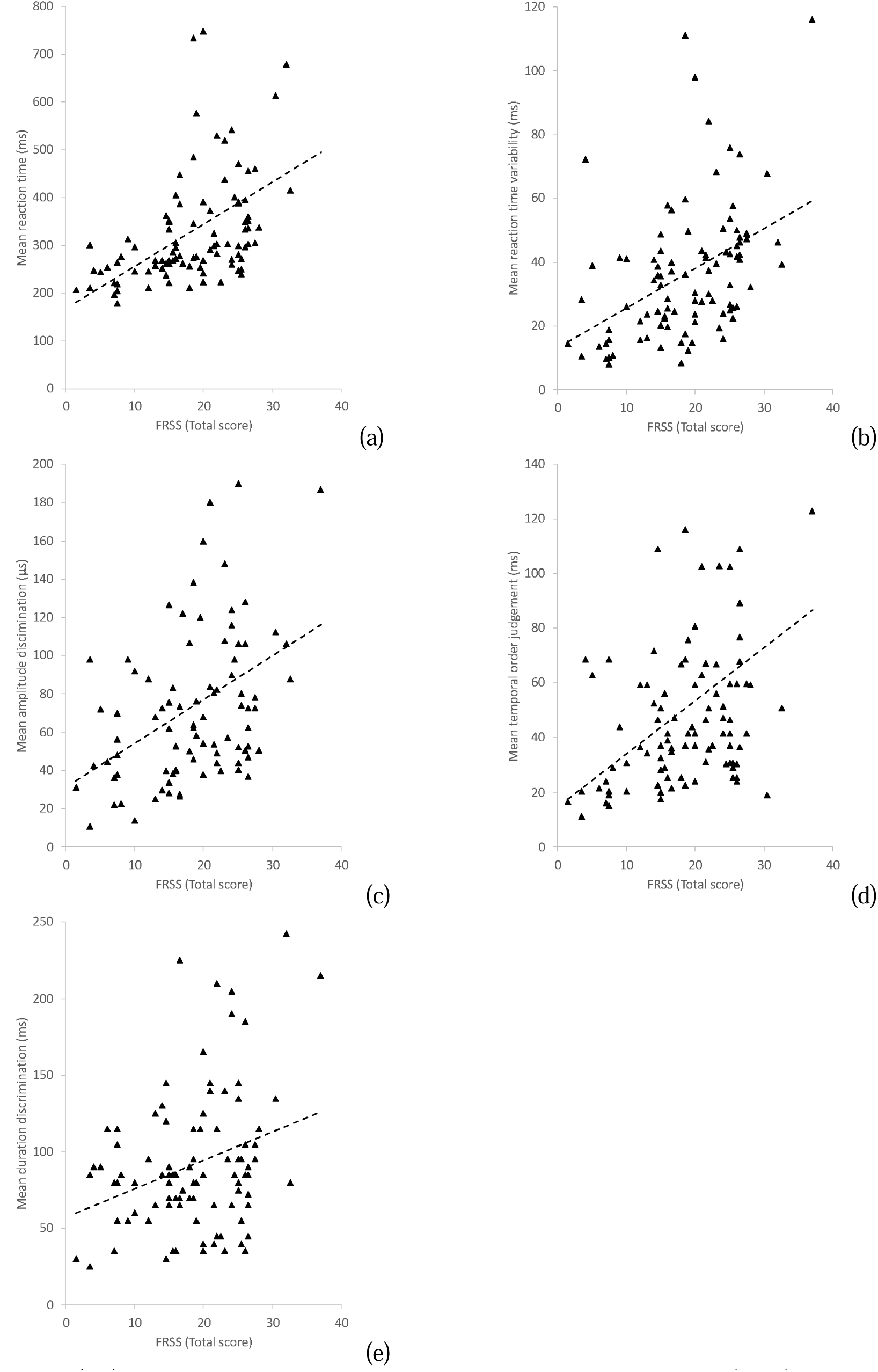
**(a–e).** Correlations comparisons between fatigue and related symptom scores (FRSS) and raw data in all participants by (a) mean Reaction Time; (b) mean Reaction Time Variability; (c) Mean Amplitude Discrimination; (d) mean Temporal order Judgement; and (e) mean Duration Discrimination.

Significant correlations were also observed between total exposure years played and TOJ (rho=0.244; P=0.025), and RT2 (rho=0.273; P=0.011) only. No significant correlations were found between professional career length and somatosensory variables.

### Correlations: comparison between groups

Comparisons of correlations between symptomatic and non-symptomatic groups (**Figure 2a – e**) revealed significant correlations in the symptomatic group between fatigue and related symptoms scores and mean RT (rho=0.309, P=0.014; **figure 2a**), mean RTvar (rho=0.291, P=0.021; **figure 2b**), mean AD (rho=0.277, P=0.028; **figure 2c**); TOJ (rho=0.273, P=0.031; **figure 2d**) and DD (rho=0.260, P=0.040; **figure 2e**). Conversely, apart from TOJ, the non-symptomatic group showed no significant correlations between fatigue and related symptoms scores and variables (mean RT: rho=0.283, P=0.203, **figure 2a**; mean RTvar: rho=0.287, P=0.196, **figure 2b**; mean AD: rho=0.154, P=0.493, **figure 2c**; TOJ: rho=0.352, P=0.010, **figure 2d**; and DD: rho=0.283, P=0.203, **figure 2e**). However, comparisons of correlations between groups were not significantly different (mean RT: Z=0.431, P=0.666; mean RTvar: Z=0.384, P=0.701; mean AD: Z=0.491, P=0.623; DD: Z=0.335, P=0.737; TOJ: Z=-1.110, P=0.267).

**Figure 2.**
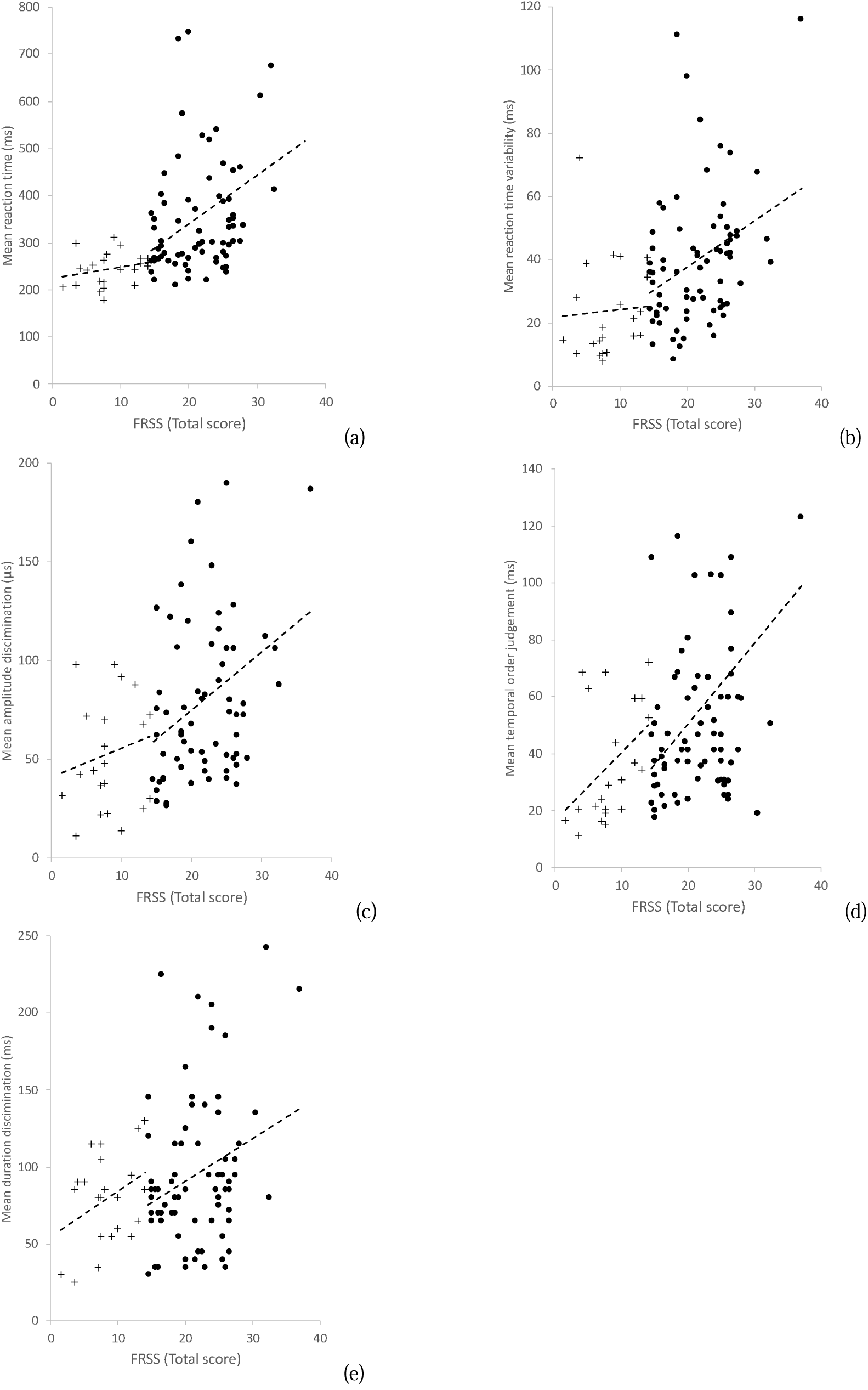
**(a – e).** Correlations comparisons between fatigue and related symptom scores (FRSS) and raw data between symptomatic (circles) and non-symptomatic (cross) groups for (a) mean Reaction Time; (b) mean Reaction Time Variability; (c) Mean Amplitude Discrimination; (d) mean Temporal order Judgement; (e) mean Duration Discrimination.

## Discussion

This study investigated the effect of exposure and concussion history on the somatosensory system in retired contact professional athletes. Between group comparisons supported our primary hypothesis, with significant differences and moderate to large effects across the majority of the variables tested. The exception was DD (although small-to-moderate effect size differences were observed). Further, correlations showed strong associations with severity of symptom reporting and somatosensory performance, with stronger correlations seen in the symptomatic group compared to the non-symptomatic group. However, comparing correlations between groups showed no differences. Results partially supported our secondary hypothesis that total exposure years was associated with TOJ and repeated testing of simple RT. No associations were found with somatosensory data and the number of concussions diagnosed.

Our findings illustrate that retired athletes who reported ongoing symptoms showed significantly higher and more variable RTs, reflecting slower perceptual-motor processing. We have previously shown increased RTs and greater variability in RTs in those with persistent post-concussion symptoms (PPCS) (Pearce et al. 2020). In those with PPCS, it is posited that chronic glial inflammation may be a contributing factor (Johansson and Rönnbäck 2014). However, in this study, participants were well beyond the acknowledged time period for PPCS. Rather we speculate the slowed and greater variability in reaction time may be a maladaptive neuroplastic response affecting information processing between brain regions applicable for vibrotactile perception and accompanying motor preparation and execution. Evidence for this come from previous TMS investigations in retired athletes who, compared to age-matched controls showed impaired neuroplastic changes through alterations in corticomotor inhibition, despite their last reported concussions were two decades prior (De Beaumont et al. 2009; Pearce et al. 2014; Pearce et al. 2018; Pearce et al. 2021; Pearce et al. 2023). Our results also showed altered somatosensory perception indicating supporting TMS cortical inhibition changes. Specifically, those who reported their total fatigue and related symptoms score >14 showed significantly worse overall AD scores as well as poorer TOJ performance (all moderate-to-large effects). While not significant (P=0.429), we observed a small-to-moderate effect (d=0.3) between groups in DD.

Higher discrimination thresholds in the symptomatic group suggest a reduced ability to discriminate spatially distinct stimuli, which has been linked to GABA-mediated lateral cortical inhibition (Tommerdahl et al. 2010; Puts et al. 2011). While GABA was not specifically measured in this study, support for disrupted GABA comes from related neurophysiological studies of chronic effects of repeated concussions and neurotrauma in both TMS (De Beaumont et al. 2009; Pearce et al. 2014; Pearce et al. 2018; Pearce et al. 2021; Pearce et al. 2023) and magnetoencephalography (Huang et al. 2020) studies.

Collectively these techniques (somatosensory and TMS) are able to provide sensitive biomarkers for detection of abnormalities and have the potential to assist clinical diagnostic criteria, currently in development, for those suspected of neurodegenerative diseases associated with repetitive neurotrauma (Katz et al. 2021).

Emerging evidence from neuropathology and neurophysiology studies are suggesting that long term or chronic neurological concerns associated with contact sports is from total exposure risk, rather than the history of concussion injuries (Daneshvar et al. 2023; Stewart et al. 2023; Pearce et al. 2024). While we did not find any correlations between the number of reported concussions and somatosensory results, we were surprised that only several significant correlations were found with regards to total exposure time and somatosensory data. Specifically, significant moderate correlations were observed only in TOJ, reflecting processing difficulties between fronto-striatal pathways (Zhang et al. 2011; Favorov et al. 2019). Moderate correlations were also observed in simple RT at the end of the test battery (RT2) suggesting increased fatigue effects, which has been a key finding in these cohorts in TMS studies (Pearce et al. 2021).

Conversely, moderate-to-strong significant correlations were observed between symptom scale total scores and somatosensory results. While the fatigue and related symptom survey is self-reported in nature, the high reliability of the scale (Chronbach’s α = 0.94 (Johansson et al. 2009)) gives us confidence that participants did not exaggerate or were careless with their responses (Ward and Meade 2023). Indeed, the somatosensory results suggests that objective testing methods such as this could be useful in mitigating concerns regarding survey methods.

While group differences were detected for all but one metric (DD), another limitation of this study relates to the small non-symptomatic group. Difficulties, and hence justified concerns around ‘sampling bias’, arise when retired professional athletes who claim to be unaffected (non-symptomatic) were less likely to respond to recruitment announcements.

However, considering disparate groups sizes, we aimed to meet minimum sample size of 83 (power 1-β err prob) based upon a group split of ∼75%/25% (symptomatic n=63; non-symptomatic n=22). Despite this, we still caution against generalisation in light of these small sample size limitations. Further work will incorporate non-elite retired athletes which will allow for greater sampler sizes.

A final limitation of this study involves the presentation of data at group levels making clinical translation to practice difficult. As long term concerns in athletes with a history of neurotrauma tend to be heterogenous (McKee et al. 2014) individual specific differences are lost within group data presentation. However, it is not possible to look at individual results without prior specific individual clinical histories, rather than a symptom self-report and retrospective recall on playing histories. Future prospective designs where concussion history and clinical data is recorded will strengthen current studies requiring retrospective data. With the rise in women’s contact sports participation, the opportunities for long-term studies in female athletes is also required.

In conclusion, this study utilised a novel approach to assessed cortical physiology in retired professional athletes using the somatosensory system. Findings showed altered sensory perception related to aberrant cortical inhibitory function in those with ongoing symptoms affecting daily activities. Further, correlations showed associations with overall severity of symptoms affecting daily activities and somatosensory performance. However, correlations did not discriminate between those symptomatic and non-symptomatic. This study is an important step expanding on pathophysiological data associated with repetitive neurotrauma demonstrating the feasibility of quantitatively assessing somatosensory perception in retired athletes. It provides a relatively low-cost clinical tool to objectively assess those who report concerns with a history of repeated brain injuries.

## Data Availability

All data produced in the present study are available upon reasonable request to the corresponding author.

